# Interleukin 1 beta is associated with depression in Multiple Sclerosis patients

**DOI:** 10.1101/2020.12.11.20245472

**Authors:** Hugo Aguilar-Talamantes, Azul Islas-Hernandez, Aleida Rodriguez-Castañenda, Brenda Bertado-Cortés, Irma Corlay-Noriega, Paola Garcia-delaTorre

## Abstract

Depressive and anxiety symptoms occur more frequently in chronic encephalomyelitis. Inflammatory diseases are highly associated with psychiatric comorbidities, which has been well established in Multiple Sclerosis. However, no biomarkers have been found with the capacity to discern between MS and depression. Thirty-six individuals with a diagnosis of MS according to the revised McDonald criteria, were recruited from an outpatient Neurology and Psychiatry from the medical unit of high specialty in Mexico. We measured the association between BDNF, IL-1β, and TNFα in serum with the presence of depression and anxiety using the semi-structured psychiatric interview, the Beck Depression Inventory (IDB), and Hospital Anxiety and Depression Scale (HADS). This was a cross-sectional study. The Logistic Regression was used for the multivariate analysis adjusted by the Multiple Sclerosis Severity Score (MSSS). With a power of 0.75 in the final model, patients with multiple sclerosis, depression, and anxiety obtained the highest values of IL-1 β in our study.

## Introduction

Multiple Sclerosis (MS) is an autoimmune disease characterized by the destruction of myelin in the central nervous system (CNS) following a pathologic triad of inflammation, demyelination, and gliosis. It mainly affects young adults, and its disease course includes recurrence-remission or progression. Lesions occur at different times and places of the CNS, and manifestations of the disease vary from a benign course to a disabling condition with fast and aggressive evolution, that obliges major lifestyle modifications for the patient [1, 2].

Inflammatory diseases are highly associated with psychiatric comorbidities, which has been well established in Multiple Sclerosis (MS). A systematic review of comorbidities in MS patients showed a prevalence of 21.9% comorbidity with anxiety, 23.7% with depression, and 4.3% with psychosis [3]. In Mexico, according to the REMEMBer study, 12% of MS patients presented comorbidity with anxiety while 17.7% with depression [4].

Depression and anxiety have a significant impact on MS patients which is associated with a reduction in their overall quality of life. Still, the treatment for these comorbidities in patients with MS is limited due to a lack of validated detection tools that allow identifying these conditions given the fact that many of the diagnostic items of depression scales are affected by the symptoms of the disease itself [5].

In this regard, Major Depressive Disorder (MDD) is diagnosed after a semistructured interview that includes clinical symptoms (fatigue, drowsiness, anxiety, sadness, anhedonia, lack of concentration, suicidal thoughts), the natural history of the disease, risk factors that could give origin to the disease and congruence to the mental test [6]; consequently, the diagnosis of MDD is merely clinical.

It has been found that patients with major depression who do not present comorbidity with any other disease have activated inflammatory pathways, including an increase in inflammatory cytokines and their soluble receptors in peripheral blood and cerebrospinal fluid. Among the most frequently described are interleukin (IL)-6, C-reactive protein, IL-1-β, and tumor necrosis factor (TNF) –α [7]. Conversely, there appears to be a decrease in brain-derived neurotrophic factor (BDNF) levels in depressed patients, which are known to increase as a consequence of antidepressant treatment [8].

To make a more accurate diagnosis of depression in MS patients, it is necessary to take into account the changes that occur in the pro-inflammatory pathways as well as in the BDNF levels for both diseases [9]. Hence, in this study, we set out to evaluate the possible correlation between the presence of depressive symptoms with an increase in pro-inflammatory cytokines and how this befalls in multiple sclerosis.

## Methods

### Study

A convenience sample was used, all subjects were recruited between September 2012 and April 2014. Prospective recruitment of all participants was done in the Psychiatry Service of the Specialties Hospital in Centro Médico Nacional Siglo XXI of the Instituto Mexicano del Seguro Social, Mexico City, Mexico. This study was approved by the Local Research and Ethics in Health Research Comities of *Hospital de Especialidades Dr. Bernardo Sepúlveda Gutiérrez* (17CI09015034) with the registration number R-2017-3601-71.

We used an observational cause-effect, ambispective study design.

### Participants

All participants agreed to be included in this study and signed a letter of informed consent. Inclusion criteria: subjects with multiple sclerosis diagnosis according to McDonald 2010, older than 18 and younger than 55 years, with any treatment.

Exclusion criteria: Subjects with any other autoimmune pathology.

### Consort diagram

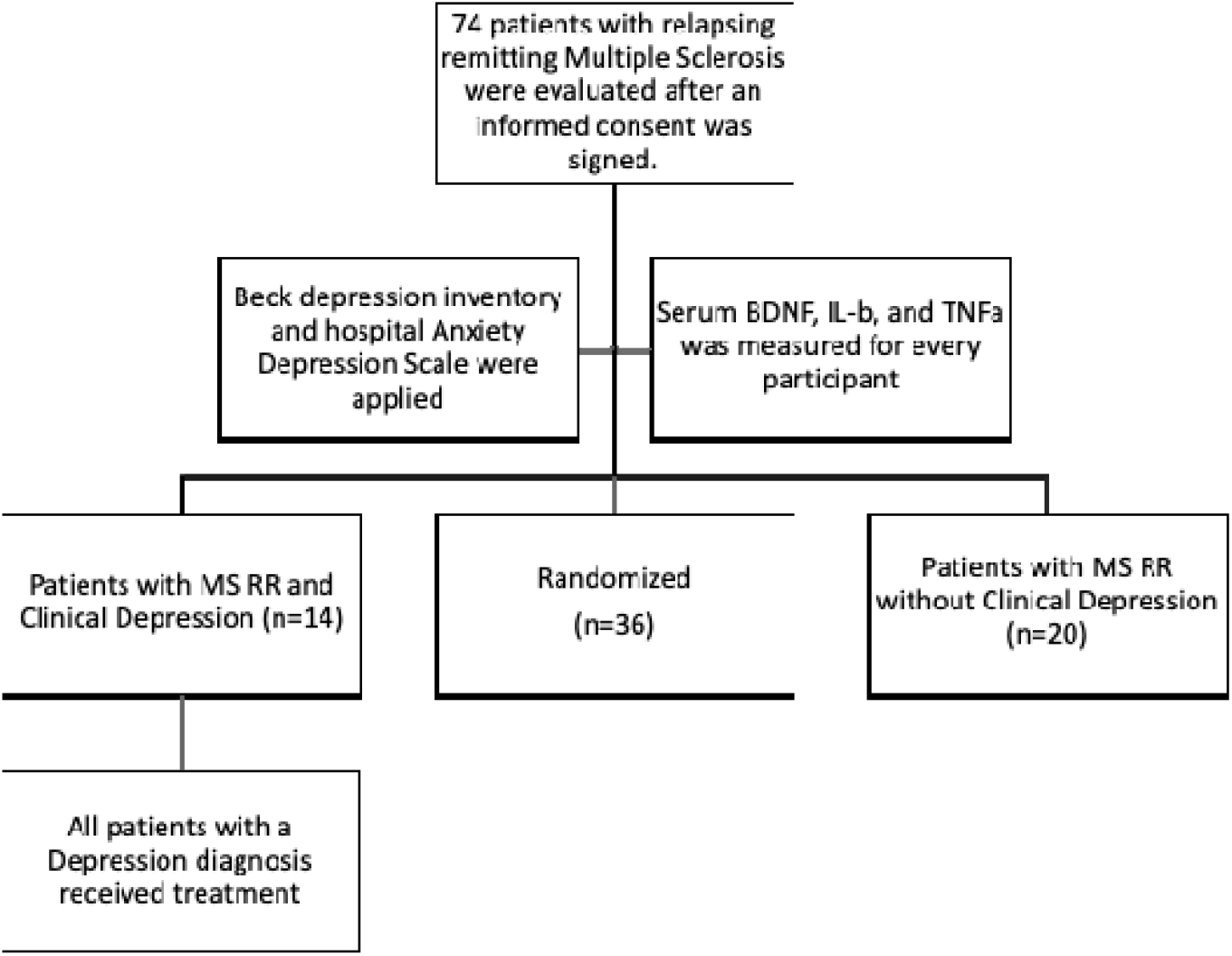

### Western Blot

Blood was collected in anticoagulant-free tubes and centrifuged at 2000g for 10min; serum was collected, and samples were stored at −40ºC for further use. No more than 2 months after the sample was taken a standard Western blotting analysis was performed to detect BDNF, IL-1B, and TNFa levels in serum.

Protein concentration was measured using the DCTM Protein Assay Kit (BioRad) according to the manufacturer’s suggestions. Sixty μg of protein per sample was separated by 14% sodium dodecyl sulfate-polyacrylamide gel electrophoresis. Samples were electrotransferred onto a polyvinylidene fluoride membrane (BioRad) and subsequently blocked with 3% skim milk in TBS-T at 4ºC overnight. The membranes were then incubated 120 min at room temperature with rabbit polyclonal antibody against BDNF (Santa Cruz; sc-5486; 1:1000 dilution), IL-1b (Santa Cruz; sc-52012; 1:200), TNFa (Santa Cruz; sc-1349; 1:500), and rabbit monoclonal antibody against Transferrin used as an internal control (Santa Cruz; sc-30159; 1:500 dilution). Blots were then incubated with horseradish peroxidase-conjugated goat anti-rabbit IgG (1:20000; PI-1000, Vector Labs, USA) in TBS-T, for 2 hrs at room temperature.

Every sample was run and analyzed twice. Protein bands were identified using chemiluminescence (LuminataTM Classico Western HRP Substrate). Images were collected using a Fusion FX imaging system (Vilber Lourmat, Marne-la-Vallée, France) and measured by densitometry using ImageJ software. Transferrin was used as a normalization control protein. For each sample, the relative abundance of the protein of interest was determined by calculating the ratio of the intensity of the signal for the protein of interest to that of the normalization control. Then, an average of both measurements per sample was obtained and used for statistical analysis.

### Ethics statement

All work was conducted with the formal approval of the human subject committees. The ethical and scientific committees of the Instituto Mexicano del Seguro Social (study number R-2017-3601-71) approved the study. This study was performed according to the World Medical Association Declaration of Helsinki.

All patients gave written consent for the study, including the acknowledgment of full anonymity. Mandatory health and safety procedures have been followed in the course of the study.

### Statistical analysis

Normal distribution was found for serum values of BDNF, IL-1β, and TNFα through the non-parametric test and the z distribution to know the mean proportion and its difference depending on if patients suffer or not depression or anxiety. We used the 75th percentile and Spearman test to select the best cutoff points of serum variables. We used the chi-square test and 2×2 tables to prove a significant difference depending on the presence or absence of depression or anxiety.

In order to evaluate the differences and associations between psychiatric symptoms and the Multiple Sclerosis Severity Score, we used a chi-square test, Spearman correlation test, and 2×2 tables.

Finally, we employed a binary logistic regression to identify the ORP of serum variables in relation to clinical diagnosis of depression, adjusting its values with MSSS (Multiple Sclerosis Severity Score) and the presence of significant symptoms of anxiety through HADS (Hospital Anxiety and Depression Scale).

## Results

A sample size of 34 patients with Multiple Sclerosis in remitting episodes was obtained; 44% were treated with Glatiramer Acetate, while 66% received some other treatment (Fingolimod, Natalizumab, Ocrelizumab, Rituximab). Table 1 shows the demographics of the population.

**Table 1.** Demographics.

| Table 1. Demographics |  |  |  |
| --- | --- | --- | --- |
|  | Female | Male |  |
| Gender | 23 (68%) | 11 (32%) |  |
| Age | 35.8 (SD 9.7) |  |  |
|  | No | Yes |  |
| Clinical Diagnosis of Depression | 20 (59%) | 14 (41%) |  |
|  | Percentile |  |  |
|  | 25 | 50 | 75 |
| Beck Depression Inventory | 6.75 | 11 | 16.25 |
| HADS Total | 7.75 | 12 | 16.75 |
| Depression subscale | 3 | 5.5 | 9 |
| Anxiety subscale | 3 | 6 | 9.5 |
| EDSS | 1 | 1 | 5 |
| MSSS | 1.45 | 2.44 | 6.53 |

When analyzing differences in biomarkers levels according to the clinical evaluations, a normal distribution of the data from the three biomarkers analyzed was found with the Kolmogorov-Smirnov test. Using a Student t-test we found significant differences of IL-1b levels according to the clinical diagnosis of depression, but no differences for BDNF or TNFa (Table 2). A cut-off point of 1.4 in the IL-1b/transferrin ratio was established using the descriptive analysis of the data.

**Table 2.** Distribution and clinical comparison of the biomarkers in Multiple Sclerosis patients.

|  |  |  |  |  |  |  |
| --- | --- | --- | --- | --- | --- | --- |
| Variables | Average Percentile. | EDSS | MSSS | Clinical diagnosis of Depression. | IDB | HADS. |
|  | (SD) 25-50-75. |  |  |  |  |  |
| BDNF | 1.4<br>1.07-1.24-1.66 | 0.17 | 0.17 | 0.5 | 0.8 | 0.4 |
|  | -0.66 |  |  |  |  |  |
| IL-1b | 1.28<br>0.61-1.06-1.48 | 0.74 | 0.74 | <u>0.04</u> | 0.41 | 0.17 |
|  | -0.93 |  |  |  |  |  |
| TNFa | 1.16<br>0.96-1.06-1.35 | 0.32 | 0.32 | 0.9 | 0.54 | 0.28 |
|  | -0.34 |  |  |  |  |  |
| Expanded Disability Status Scale (EDSS). Multiple Sclerosis Severity Score (MSSS). Beck Depression Inventory (IDB). Hospital Anxiety and Depression Scale (HADS). |  |  |  |  |  |  |

When dichotomizing EDSS (EDSS < 3 = 0, 3 or more = 1) and MSSS (MSSS < 4 = 0, 4 o more = 1), both with the same proportion, we found the same statistical differences for both scales for each biomarker, none of which were significant (Table 2); showing that both dichotomized measurements behave the same way.

We then evaluated the differences and association between psychiatric symptoms and the Multiple Sclerosis Severity Score. We found differences using a Chi-squared test for HADS >7 and IDB >9 as well as associations, using Spearman’s rank correlation, with both scores used to evaluate depression (IDB and HADS) and the Multiple Sclerosis Severity Score. A higher decile in the MSSS is associated with depression (higher IBD score) and anxiety (a higher HADS score), conferring a higher severity of MS as a factor for comorbidity with these symptoms (Table 3). Therefore, the probability of testing positive for depression using either test was 5 to 6 times greater when the severity of Multiple Sclerosis was of the fourth decile or greater.

**Table 3.**
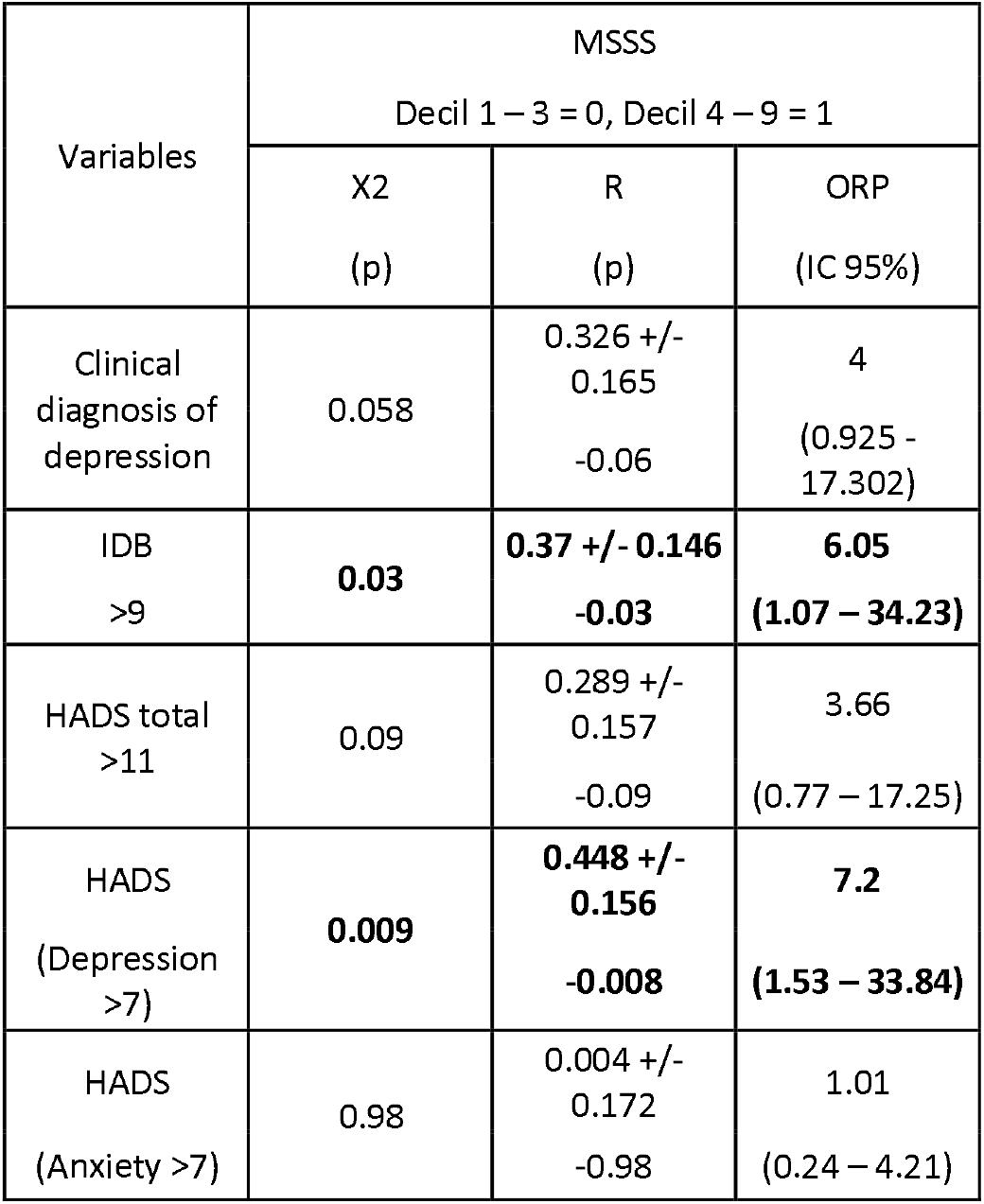
Differences and association between psychiatric symptoms and Multiple Sclerosis Severity Score.

When the clinical variables (MSSS and anxiety symptoms) and the biomarkers were analyzed, it was possible to observe that values of IL-1b > 1.4 are more frequent in patients with clinical depression (Figure 1).

When we tested the variables dichotomized in the Logistic Regression, we obtained an ORP of 28.5 for clinical diagnosis of depression and an ORP of 8.6 and 3.1 for the Beck Depression Inventory in its two cut-off points for diagnosis of depression (>14 and >9 respectively). As a result, our model had its best performance in the association with the clinical diagnosis of depression. Table 4 shows that the model allows an estimation of 100% for the probability of ruling out a psychopathological process in patients with Multiple Sclerosis in Remission.

**Table 4.** Binary Logistic Model for the clinical diagnosis of depression in patients with Multiple Sclerosis.

|  |  |  |  |  |  |  |  |
| --- | --- | --- | --- | --- | --- | --- | --- |
| Variables in the Model | Value of B | R<sup>2</sup><br>Nagelkerke | ORP<br>(IC 95%) | Sensitivity | Specificity | Positive Predictive Value | Negative Predictive Value |
| MSSS >4th decile | 2.695 +/- 1.337 | 0.616 | 28.503<br>(2.166 – 375.034) | 100% | 80% | 64% | 100% |
| IL-1b/Transferrin >1.400 | 2.263 +/- 1.332 |  |  |  |  |  |  |
| HADS anxiety >7 | 3.350 +/- 1.315 |  |  |  |  |  |  |
| Constant | -3.392 +/- 1.25 |  |  |  |  |  |  |

## Discussion

During the progression of MS, axonal demyelination causes a constant state of inflammation in the brain, exacerbated by the activation of microglia [10]. The anxiety and depression that occur during MS have been considered a consequence of the awareness that the patient has of the disease; however, it is now known that their relationship is more complex, pointing to inflammatory factors typical of MS and the presence of depressive and/or anxiety symptoms.

In this study, we found a 100% estimation for the probability of ruling out a psychopathological process in MS with Remission patients when the MSSS was < 4, with low anxiety symptoms (HADS, the subscale of anxiety < 7) and serum levels of IL-1b < 1.4 measured by western blot analysis.

Using a statistical multivariate model, we found that, when we have a result <4 on the MSSS scale, psychopathological processes can be ruled out in patients with MS with remission, with low levels of anxiety (HADS, the subscale of anxiety <7) and with low levels of IL-1b in serum.

The golden standard for depression diagnosis relies on a personal semi-structured interview using DSM5 criteria ([6]). No biomarkers are currently used for the clinical diagnosis of depression with or without comorbidities even though several molecules have been found in experimental studies [11–13]. Finding a biomarker that can be used independently of the comorbidity is essential and extremely difficult since depression is commonly found simultaneously with other medical conditions [14].

Other studies have found an association between inflammatory biomarkers and depression in comorbidity. However, most of these studies do not use the DSM5 criteria nor the gold standard for the diagnosis of depression [15–17] and only use depression symptoms, this could explain the low association margin obtained for depression and the given comorbidities. Here we found a huge gap of association when using the clinical diagnosis (ORP=28.5) or the Beck Depression Inventory with two cut-off points (ORP=8.6 and ORP=3.1). In order to find a proper biomarker for depression, the clinical diagnosis must be taken into account.

We found that IL-1b can be used to discriminate psychopathology (depression and/or anxiety) from Multiple Sclerosis. Ruling out this comorbidity would allow the clinicians to infer a favorable short-term prognostic as has been seen in studies where an association of depression with the worst outcome has been seen [18].

Regarding IL-1b, it is known that its administration, whether systemic or central, causes depressive-type symptoms in animals, and it has been speculated that this is because cytokines have an effect on glutamatergic transmission, as well as on the function of various neurotransmitters [19].

On that matter, IL-1b is also the link between the immune system and the CNS [20], it modulates the metabolism of tryptophan by increasing the activity of 2,3-IDO, generating a lower availability of tryptophan for the production of serotonin, and indirectly increasing the production of potentially neurotoxic metabolites [21]. Increased central IL-1b activity has also been reported to contribute to cortisol resistance seen in several cases of patients with depression. It can also influence neuronal plasticity through the activation of microglia [22].

On the other hand, we did not find an association between TNFa, a pleiotropic cytokine that plays an important factor in the early stages of the inflammatory response [23], and the presence of depression and/or anxiety. On the matter, when TNFa was systemically administered in rodents they found that although they showed signs of sickness-like behavior (body weight loss and a reduced locomotor activity), none showed depressive or anxious symptoms (anhedonia), concluding that the increase in TNFa is not directly related to this type of symptoms [24]. Also, one of our inclusion criteria was not being treated with interferon-beta because it is an independent risk factor for depression and anxiety in patients with multiple sclerosis [25].

As with other comorbidities, depression in MS is normally ignored. For example, the Expanded Disability Status Scale (EDSS) gives a minimal value to depression which by itself does not affect the EDSS punctuation and is merely mentioned as part of other symptoms related to MS. This exclusion affects the patient in more than one aspect. A proper diagnosis of depression would allow proper treatment, and as mentioned before, depression affects prognosis in diseases such as Rheumatoid Arthritis [26], Chronic Kidney Disease [27], type 2 diabetes [28], irritable bowel syndrome [29], and Multiple Sclerosis [18].

One of the main issues with this study is the sample size, nonetheless, the statistical power (P=0.8) was enough to conclude that the diagnosis of depression can use biomarkers such as IL-1b as part of its clinical characterization even when comorbidity such as MS is present, with the possibility of conjointly using other clinical variables to exclude the possibility of a psychopathological process with a P=0.7. Other comorbidities with depression need to be evaluated in order to achieve a real biological diagnosis that can accompany the Gold Standard in Psychiatry.

A follow up of the patients that present these risk factors (MSSS was > 4, HADS subscale of anxiety > 7, and levels > 1.4 in the serum values of IL-1b) would be advisable in order to identify if these variables are also associated with a worst prognosis of MS due to the presence of inflammatory depression.

Inflammation caused by the presence of MS can induce depressive symptoms. Inflammation is, therefore, a biological event, which must be taken into account alongside the traditional psychosocial factors implicated in this kind of mood disorder. Using serum IL-1b measurements we obtained a 100% estimation for the probability of ruling out a psychopathological process in MS with Remission patients. A logistic model where MSSS was < 4, HADS subscale of anxiety was < 7, and serum IL-1b values were < 1.4 allowed a clinical approximation for the use of biomarkers in depression diagnosis.

## Supporting information

STROBE

## Data Availability

The authors confirm that the data supporting the findings of this study are available within the article.

## Acknowledgments

The grant FIS/IMSS/PROT/G12/1129 was used to fulfill this study. There was no involvement of the founder in the elaboration or analysis of this study.

## References

1. Wiener C, Fauci AS, Braunwald E, Kasper DL, Hauser S, Longo D, et al. Harrisons Principles of Internal Medicine Self-Assessment and Board Review. McGraw Hill Professional; 2016.

2. Ropper A, Samuels M, Klein J. Adams, and Victor’s Principles of Neurology 10th Edition. McGraw Hill Professional; 2014.

3. Marrie RA, Reingold S, Cohen J, Stuve O, Trojano M, Sorensen PS, et al. The incidence and prevalence of psychiatric disorders in multiple sclerosis: A systematic review. Multiple Sclerosis Journal. 2015;21:305–317.

4. Bertado-Cortés B, Venzor-Mendoza C, Rubio-Ordoñez D, Pérez-Pérez JR, Novelo-Manzano LA, Villamil-Osorio LV, et al. Demographic and clinical characterization of multiple sclerosis in Mexico: The REMEMBer study. Multiple Sclerosis and Related Disorders. 2020;46:102575.

5. Watson TM, Ford E, Worthington E, Lincoln NB. Validation of mood measures for people with multiple sclerosis. Int J MS Care. 2014;16:105–109.

6. American Psychiatric Association. Diagnostic and Statistical Manual of Mental Disorders (DSM-5^®^). American Psychiatric Pub; 2013.

7. Raison CL, Capuron L, Miller AH. Cytokines sing the blues: inflammation and the pathogenesis of depression. Trends Immunol. 2006;27:24–31.

8. Więdłocha M, Marcinowicz P, Krupa R, Janoska-Jaździk M, Janus M, Dębowska W, et al. Effect of antidepressant treatment on peripheral inflammation markers - A meta-analysis. Prog Neuropsychopharmacol Biol Psychiatry. 2018;80:217–226.

9. Lopresti AL, Hood SD, Drummond PD. A review of lifestyle factors that contribute to important pathways associated with major depression: diet, sleep, and exercise. J Affect Disord. 2013;148:12–27.

10. Correale J, Marrodan M, Ysrraelit MC. Mechanisms of Neurodegeneration and Axonal Dysfunction in Progressive Multiple Sclerosis. Biomedicines. 2019;7.

11. Lopresti AL, Drummond PD. Obesity and psychiatric disorders: commonalities in dysregulated biological pathways and their implications for treatment. Prog Neuropsychopharmacol Biol Psychiatry. 2013;45:92–99.

12. Strawbridge R, Arnone D, Danese A, Papadopoulos A, Herane Vives A, Cleare AJ. Inflammation and clinical response to treatment in depression: A meta-analysis. Eur Neuropsychopharmacol. 2015;25:1532–1543.

13. Sen S, Duman R, Sanacora G. Serum brain-derived neurotrophic factor, depression, and antidepressant medications: meta-analyses and implications. Biol Psychiatry. 2008;64:527–532.

14. Kang H-J, Kim S-Y, Bae K-Y, Kim S-W, Shin I-S, Yoon J-S, et al. Comorbidity of depression with physical disorders: research and clinical implications. Chonnam Med J. 2015;51:8–18.

15. Baghai TC, Varallo-Bedarida G, Born C, Häfner S, Schüle C, Eser D, et al. Classical Risk Factors and Inflammatory Biomarkers: One of the Missing Biological Links between Cardiovascular Disease and Major Depressive Disorder. Int J Mol Sci. 2018;19.

16. Cheon Y-H, Lee S-G, Kim M, Kim H-O, Sun Suh Y, Park K-S, et al. The association of disease activity, pro-inflammatory cytokines, and neurotrophic factors with depression in patients with rheumatoid arthritis. Brain Behav Immun. 2018;73:274–281.

17. Geng Q, Zhang Q-E, Wang F, Zheng W, Ng CH, Ungvari GS, et al. Corrigendum to ‘Comparison of comorbid depression between irritable bowel syndrome and inflammatory bowel disease: A meta-analysis of comparative studies’ [J Affect Disord. 2018 237: 37-46. DOI: 10.1016/j.jad.2018.04.111.]. J Affect Disord. 2019;253.

18. Fiest KM, Walker JR, Bernstein CN, Graff LA, Zarychanski R, Abou-Setta AM, et al. Systematic review and meta-analysis of interventions for depression and anxiety in persons with multiple sclerosis. Mult Scler Relat Disord. 2016;5:12–26.

19. Dantzer R. Cytokine-induced sickness behavior: where do we stand? Brain Behav Immun. 2001;15:7–24.

20. Baganz NL, Blakely RD. A dialogue between the immune system and brain, spoken in the language of serotonin. ACS Chem Neurosci. 2013;4:48–63.

21. Dantzer R, O’Connor JC, Lawson MA, Kelley KW. Inflammation-associated depression: From serotonin to kynurenine. Psychoneuroendocrinology. 2011;36:426–436.

22. Miller AH, Maletic V, Raison CL. Inflammation and Its Discontents: The Role of Cytokines in the Pathophysiology of Major Depression. Biol Psychiatry. 2009;65:732–741.

23. Clark IA, Alleva LM, Vissel B. The roles of TNF in brain dysfunction and disease. Pharmacol Ther. 2010;128:519–548.

24. Biesmans S, Bouwknecht JA, Ver Donck L, Langlois X, Acton PD, De Haes P, et al. Peripheral Administration of Tumor Necrosis Factor-Alpha Induces Neuroinflammation and Sickness but Not Depressive-Like Behavior in Mice. Biomed Res Int. 2015;2015:716920.

25. Alba Palé L, León Caballero J, Samsó Buxareu B, Salgado Serrano P, Pérez Solà V. Systematic review of depression in patients with multiple sclerosis and its relationship to interferonβ treatment. Mult Scler Relat Disord. 2017;17:138–143.

26. Cabrera-Marroquín R, Contreras-Yáñez I, Alcocer-Castillejos N, Pascual-Ramos V. Major depressive episodes are associated with poor concordance with therapy in rheumatoid arthritis patients: the impact on disease outcomes. Clin Exp Rheumatol. 2014;32:904–913.

27. Jain N, Carmody T, Minhajuddin AT, Toups M, Trivedi MH, Rush AJ, et al. Prognostic Utility of a Self-Reported Depression Questionnaire versus Clinician-Based Assessment on Renal Outcomes. Am J Nephrol. 2016;44:234–244.

28. Starkstein SE, Davis WA, Dragovic M, Cetrullo V, Davis TME, Bruce DG. Diagnostic criteria for depression in type 2 diabetes: a data-driven approach. PLoS One. 2014;9:e112049.

29. Stasi C, Caserta A, Nisita C, Cortopassi S, Fani B, Salvadori S, et al. The complex interplay between gastrointestinal and psychiatric symptoms in irritable bowel syndrome: A longitudinal assessment. J Gastroenterol Hepatol. 2019;34:713–719.

